# The impact and cost of reaching the UNAIDS global HIV targets

**DOI:** 10.1101/2025.07.01.25330647

**Authors:** John Stover, Deepak Mattur, Mariana Siapka, Triantafyllos Pliakas, Ricardo Valladares-Cardona

## Abstract

**Background:** There has been significant progress in combating the HIV/AIDS pandemic in the last 15 years. In order to describe towards epidemic control, UNAIDS convened a Global Task Team to recommend targets for 2030 for key HIV interventions. This paper describes the approach to estimate the impact and cost of achieving those targets.

**Methods and data:** We used an HIV simulation model, Goals, to simulate key epidemiological indicators through 2030 is targets are reached in 134 countries with the highest HIV burden. To estimate the cost of achieving the targets we compiled a unit cost database and applied it to estimate the cost of each intervention in each country for the years 2025 through 2030.

**Results:** Achieving the UNAIDS 2030 targets would reduce the annual number of new HIV infections globally by 77% from 2023 to 2030, from 1.3 million to 300,000. The number of AIDS deaths would decline by 50% over the same period. The number of people living with HIV would decline slightly, from 40 million to 39 million, but would continue declining to 35 million by 2040. The number of people receiving ART would increase from about 31 million to almost 36 million by 2030 before declining to 32 million by 2040. If the targets are achieved the number of new infections would drop below the number of deaths to people living with HIV by 2026. By 2030 there would be 330,000 fewer new infections than deaths

**Discussion:** Achieving the UNAIDS targets for 2030 would reduce the annual number of new HIV infections to just 9% of the peak value in 1995 and AIDS deaths to 13% of the peak value in 2004. In 2030 new infections would be 60% less than deaths to PLHIV, setting us on a path to achieve the end of AIDS as a public health threat.

The resources required to achieve these targets are substantial but $ 5 billion less than previous estimates of $ 29.3 billion.

Several donors have announced reductions to assistance for HIV programs in 2025. Thus, the task of mobilizing these additional resources is challenging, but if it can be done, we can achieve substantial success in combating this epidemic which has affected so many lives since it began 54 years ago.

## Background

Significant progress has been made in the last 15 years in the effort to control the HIV epidemic. From 2010 to 2023 the number of new infections declined by 39% and AIDS-related deaths declined by 51%. However, in 2023 there were still 1.3 (1.0 – 1.7) million new infections and 630,000 (500,000 – 820,000) AIDS-related deaths [1].

To help coordinate the effort to end the pandemic, UNAIDS has worked with partners through a collaborative process to develop a series of global targets for key HIV interventions. These include the well-known 95-95-95 testing and treatment targets (95% of people living with HIV know their status, 95% of them are on ART and 95% of them are virally suppressed) as well as targets for programs to prevent mother-to-child transmission; provide primary prevention interventions such as condoms, pre-exposure prophylaxis (PrEP), voluntary medical male circumcision (VMMC), services for key populations; and address issues of stigma, discrimination, access to justice and gender-based violence. The last set of targets was released in 2020 [2].

In 2024 UNAIDS convened a Global Task force on 2030 Targets to update the targets for 2030. That process has recommended 16 top-line targets (Table 1) and 50 second-tier targets. The purpose of this paper is to present estimates of the impact and cost of achieving these targets.

**Table 1.** Top-line targets for 2030.

| Priority Area | 2030 Target |
| --- | --- |
| Ensure available, accessible, acceptable and quality HIV treatment and care for people living with HIV | • 95% of people living with HIV (PLHIV) know their status<br>• 95% of PLHIV who know their status receive treatment<br>• 95% of PLHIV who are on treatment have suppressed viral loads |
| Scale-up prevention options that bring together biomedical, structural and behavioral interventions | • 90% of people in need of prevention use prevention options (PrEP, post-exposure prophylaxis (PEP), condoms, VMMC, needle and syringe programs (NSP), and opioid agonist maintenance therapy (OAMT)) |
| End stigma and discrimination and uphold human rights and gender equality in the HIV response | • <10% of PLHIV or key and vulnerable populations experience stigma and discrimination<br>• <10% experience gender inequality of violence<br>• <10% of countries have punitive legal and policy environments that restrict access to services |
| Ensure community leadership in the HIV response | • Community-led organizations (CLO) deliver 30% of testing and treatment support services<br>• CLO deliver 80% of prevention options<br>• CLO delivery 60% of societal enabler programs |
| Integrate HIV services with primary health care, broader health systems and other sectors | • 95% of people who are receiving HIV prevention or treatment services also receive needed sexual and reproductive health (SRH) services (including for sexually transmitted infections (STIs))<br>• 95% of pregnant women living with HIV and their newborns receive maternal and newborn care that integrates or links to comprehensive HIV services, including for prevention of HIV and hepatitis B virus and treatment of syphilis |
| Ensure sustainable financing for a people-centered national and global HIV response | • Reduce out-of-pocket expenditures for HIV in line with Universal Health Care (UHC)<br>• Increase the percentage of HIV spending that is domestic<br>• US\$ 21.9 billion mobilized for HIV investments for low- and middle-income countries<br>• All countries have access to equitable pricing for diagnostics and therapeutics |

## Methods and data

In 2020 an HIV simulation model, Goals, was used to estimate the impact of achieving the targets [3]. We used the same approach to estimate the impact of the new targets. Goals is an HIV simulation model applied at country level to estimate the course of the HIV epidemic over time. Incidence is estimated for 11 population groups (male and females who are not sexually active, with one sexual partner in the last year, with multiple sexual partners in the last year, sex workers and their clients, people who inject drugs; and men who have sex with men) as a function of the behaviors of each group (number of partners, contacts per partner, age at first sex, needle sharing behavior) and interventions designed to male behaviors safer (services for key population, comprehensive sexuality education (CSE)) and interventions that reduce the probability of transmission per contact (condoms, PrEP, VMMC, prevention of mother-to-child transmission (PMTCT) and anti-retroviral treatment (ART)). People who become infected are followed as they progress to lower CD4 counts, get on ART or die. The model is fit to country data on HIV prevalence by population group from national population surveys, key population surveillance and antenatal clinic (ANC) testing. Goals models were calibrated for 134 countries representing 98% of all people living with HIV in the world. These models were used to compare two scenarios, one in which the current coverage of all interventions remains constant at the 2023 level and one in which the coverage of each intervention is scaled up linear from 2025 to the target value by 2030.

Estimates of the financial resources required to achieve the targets for most interventions (key population services, PrEP, NSP, OAMT, CSE, VMMC, condoms, ANC testing, early infant diagnosis, adult and pediatric ART, treatment for advanced HIV disease (AHD), testing) were based on the number of people that would receive each service at target coverage levels and the unit cost of providing each service for one year.

Resources required for programs to address gender-based violence were based on previous estimates of the needs to implement interventions with demonstrated effectiveness in all low- and middle-income countries [4]. We included 25% of global needs based on previous work on the fair share of HIV of structural interventions [5].

Resources required for programs to address stigma and discrimination were estimated based on data from a cohort of 15 countries in which costing data existed from a Global Fund Study, analyzing retrospective and prospective costing for interventions aimed at reducing stigma and discrimination. The interventions are categorized into three main areas: information, education, and communication (community awareness campaigns, and strengthening communities); empowerment programs (train the trainer, face-to-face peer training, online synchronous training, and support groups in NGOs delivering ART services.); and facility-based interventions. The cost components include start-up costs, recurrent costs, and variable costs, with assumptions made for scaling up the programs across different countries. Results extrapolated to all LMICs, variations in unit costs between countries were taken into consideration by estimating the average cost of programs across countries by income level. The study also made several assumptions for projecting program costs to 2031, including the frequency of program delivery and the intensity of scale-up to reach targets in key populations. More information is available in the Supplementary Text.

The key interventions to improve access to justice were estimated for low- and middle-income countries with high and very high HIV burden. To this end, the population of judges, police and prison guards, and prison inmates were projected, based on UNODC statistics, modelling according to income group and total population. The costs were optimized by integrating communication and information technologies in key areas: online legal support, mandatory online courses for justice operators, and social media campaigns (more effective and better targeted than mass media campaigns). The resulting figures were adjusted to intensify the investments in countries with most difficult situations of criminalization, hate crimes and violation of rights. To this end, the enabling environment index was applied.

Resources required for other services (STIs, PEP, co-morbidities, program management and above-site level costs) were estimated as a percentage of the direct intervention costs based on current spending patterns from National AIDS Spending Assessments. Ratios of support services to direct intervention spending were 1.5% for STIs, 2.0% for programs for prisoners, 0.29% for post-exposure prophylaxis, 5% for co-morbidities and 20% for program management. Resource needs for health system strengthening were set at 16% for Global Fund-eligible countries (based on Global Fund expenditure reports) and, for all other countries, a value between 0% and 16% proportional to the strength of the health system as measured by the WHO Universal health coverage service index [6].

Estimates of population sizes by age and sex are based on the United Nations’ World Population Prospects 2024 [7] and official UNAIDS estimates of the number of people on treatment and key population sizes.

Unit costs were based on published studies, estimates prepared as part of efforts to develop HIV investment cases or national strategic plans facilitated by Avenir Health, the Burnett Institute or East West Center, or modeling studies through the HIV Modeling Consortium’s MIHPSA project (Modeling to Inform HIV Programmes in Sub-Saharan Africa), WHO Last Mile collaborations and data on costing studies published on UNAIDS HIV Financial dashboard [8]. Commodity costs for ARVs, lab tests, oral PrEP, long-acting PrEP, diagnostic tests and condoms were based on current procurement prices and expert opinion of prices in 2030. These projections were informed by a technical expert consultation convened by UNAIDS, involving representatives from major donor agencies, multilateral organizations, and national governments. When country-specific values were not available regional averages of available estimates were used. S1 Table presents the unit costs by intervention and region. The full database of unit costs by intervention and country is available on GitHub [9]. The unit cost repository was reviewed and validated by national focal points, with coordination and technical facilitation provided by UNAIDS. Several countries updated national-level unit cost estimates to reflect recent procurement experiences and planning data. The unit costs for most interventions were assumed to remain constant through 2030. The exceptions were drugs for oral PrEP (which declined to $31 for low- and lower-middle income countries and $50 for all others), drugs for long- acting PrEP (which declined to $41 for low- and lower-middle income countries and $100 for all others).

Coverage targets were defined by the Global Task Team (Table 1) [10]. However, most prevention targets are based on the population who need prevention services. For services for key populations and for PrEP we defined the population in need of prevention based on incidence in that population. For key populations, incidence in 2023 was estimated by fitting the Goals model to prevalence from surveillance data. For general populations incidence by age, sex and district of residence was from UNAIDS estimates based on the Naomi geospatial model [11]. Coverage targets for 2030 by intervention are shown in Table 2.

**Table 2.** 2030 coverage targets.

| Intervention | 2030 Target |
| --- | --- |
| Services for key populations | If incidence $\geq 0.5\%$ target = 95%<br>If incidence $< 0.5\%$ target = incidence / $0.5\% \times 95\%$ |
| PrEP for FSW and PWID | If incidence $\geq 3\%$ target = 80%<br>If incidence $< 0.2\%$ target = 0%<br>If incidence between 0.2% and 3%<br>target = incidence / $(3\% - 0.2\%) \times 80\%$ |
| PrEP for MSM and TG | Target is maximum of 20% and<br>If incidence $\geq 3\%$ target = 80%<br>If incidence between 0.2% and 3%<br>target = incidence / $(3\% - 0.2\%) \times 80\%$ |
| Needle and syringe programs | If incidence $\geq 3.5\%$ trg = 95%<br>If incidence $< 3.5\%$ trg = incidence / $3.5\% \times 95\%$<br>Target = trg x PWID services coverage |
| Opioid agonist maintenance therapy | If incidence $\geq 3.5\%$ trg = 95%<br>If incidence $< 3.5\%$ trg = incidence / $3.5\% \times 95\%$<br>Target = trg x PWID services coverage |
| PrEP for male and females 15-24 and 25-49 | Targets are set by population and districts and only for countries in Sub-Saharan Africa<br>If incidence $\geq 0.2\%$ target = 80%<br>If incidence $< 0.03\%$ target = 0% |
|  | If incidence between 0.03% and 0.2% target = (incidence – 0.03%)/(0.2%-0.03%) x 80% |
| Comprehensive sexuality education | 95% of secondary school students |
| Voluntary medical male circumcision | 90% of males 15-49 in 15 priority countries (Botswana, Eswatini, Ethiopia, Kenya, Lesotho, Malawi, Mozambique, Namibia, Rwanda, South Africa, South Sudan, Tanzania, Uganda, Zambia, Zimbabwe) |
| Condoms | 80% for people who have multiple partners in the past year |
| ART | 90% of people living with HIV |
| PMTCT | 95% of HIV-positive pregnant women |
| ANC testing | 95% of women giving birth |
| Advanced HIV disease prophylaxis | 95% of people with AHD |

Note: 2030 target is always greater than or equal to reported coverage in 2023. ‘Incidence’ refers to incidence in the reference population in 2023

## Results

Achieving the UNAIDS 2030 targets would reduce the annual number of new HIV infections globally by 77% from 2023 to 2030, from 1.3 million to 300,000. The number of AIDS deaths would decline by 50% over the same period. The number of people living with HIV would decline slightly, from 40 million to 39 million, but would continue declining to 35 million by 2040. The number of people receiving ART would increase from about 31 million to almost 36 million by 2030 before declining to 32 million by 2040. If the targets are achieved the number of new infections would drop below the number of deaths to people living with HIV by 2026. By 2030 there would be 330,000 fewer new infections than deaths.

From 2010 to 2030 new infections would decline by 86% and AIDS deaths by 80%, nearly meeting the UNAIDS goal of 90% decline by 2030. (Figure 1). With constant coverage of all interventions from 2023-2030 there would be slight increases in the number of new HIV infections and AIDS deaths.

**Figure 1.**
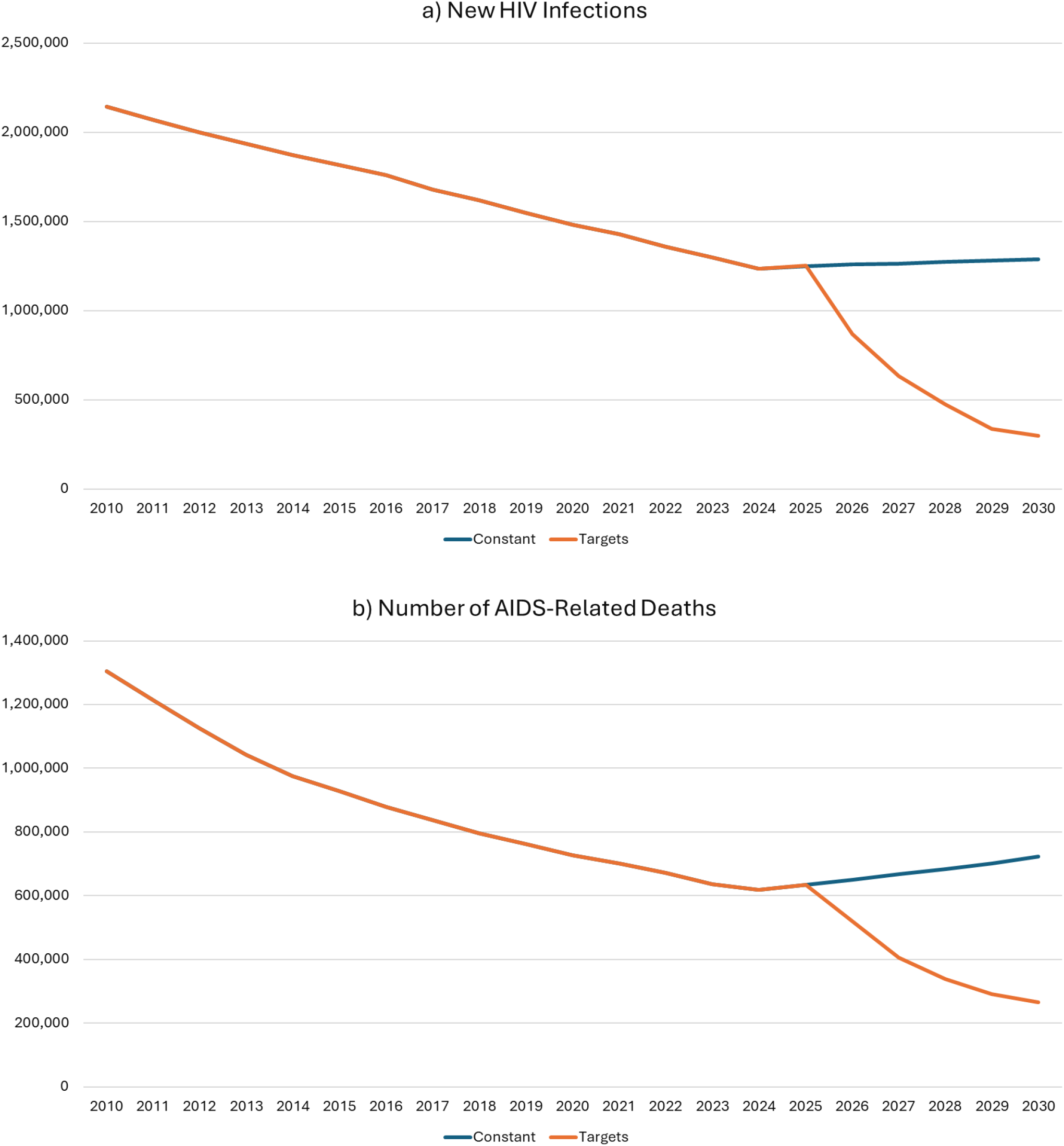
Trend in (a) new infections and (b) AIDS-related deaths with achievement of UNAIDS targets by 2030 (orange line) or constant coverage of all interventions from 2023 to 2030 (blue line). Key: KP = services for key populations except PrEP, NSP = needle and syringe programs, OAMT = opioid agonist maintenance therapy, PrEP = pre-exposure prophylaxis, CSE = comprehensive sexuality education, VMMC = voluntary medical male circumcision, ART = antiretroviral therapy, AHD = advanced HIV disease, Testing = ANC tests, EID and rapid diagnostic test, STIs = sexual transmitted infections, PEP = post-exposure prophylaxis, GBV = prevention of gender-based violence, S&D = prevention of stigma and discrimination, Justice = access to justice, HSS = health system strengthening

The largest decline in the number of new infections from 2023 to 2030 would occur in East and Southern Africa but the largest percentage declines would occur in those regions where coverage of key HIV interventions is currently low, 87% in Eastern Europe and Central Asia, 86% in North Africa and the Middle East. Similarly, the largest decline in the number of AIDS deaths would be in East and Southern Africa but the largest percentage declines would occur in Eastern Europe and Central Asia and North Africa and the Middle East. Compared to coverage remaining constant from 2023 to 2030 achieving the 2030 targets would avert 765,000 new HIV infections from 2026-2030 and 360,000 AIDS deaths.

The financial resources required to achieve these targets in low- and middle-income countries were estimated to increase to just over $21.9 billion by 2030. (Figure 2). The largest amounts for direct services are for ART (34%), prevention services for key populations (FSW, MSM, TG, PWID) except PrEP (13%), societal enabler interventions (10%). Management and health system strengthening require 15% and 10% respectively.

**Figure 2.**
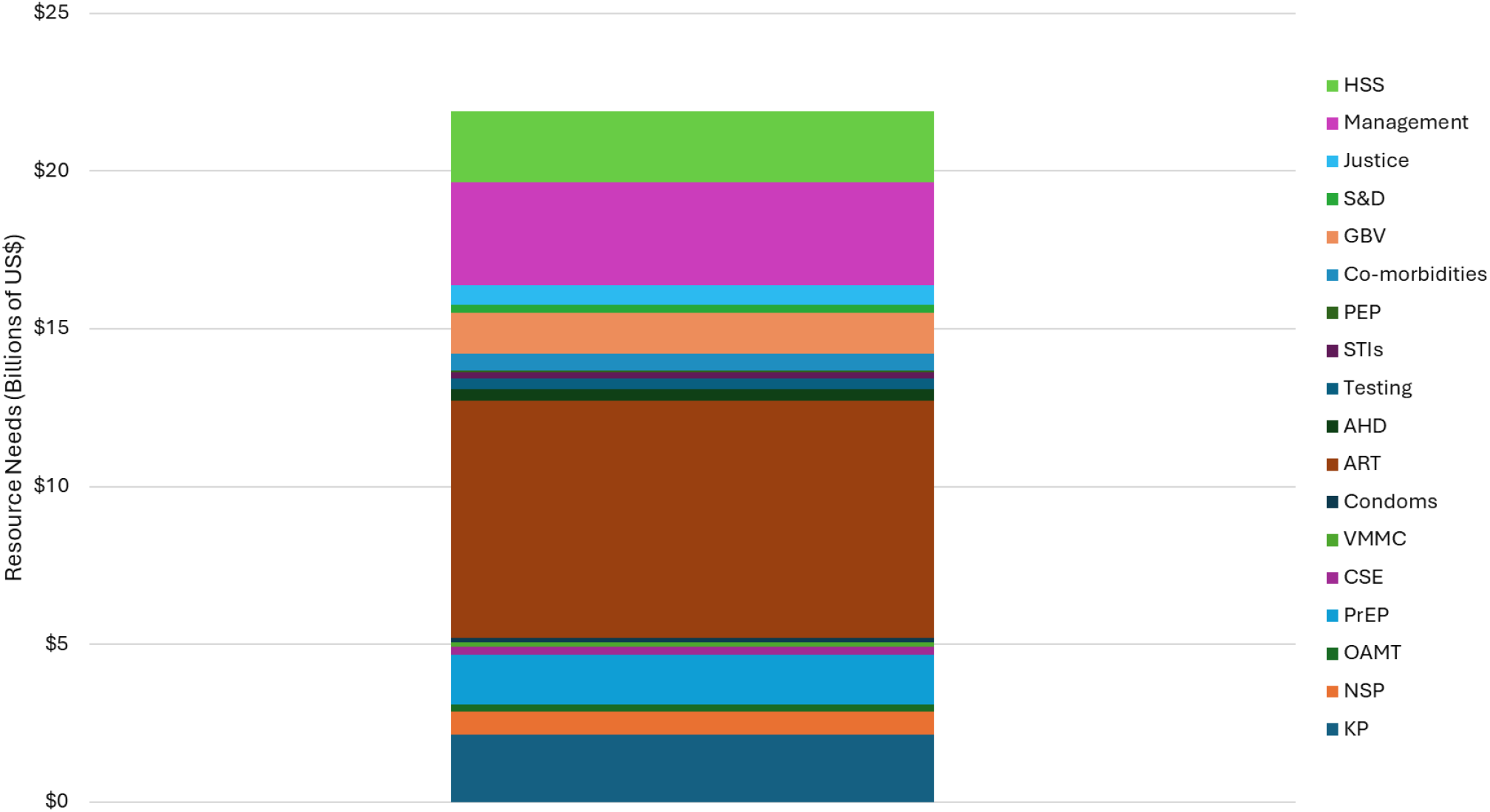
Resource needs in low- and middle-income countries in 2030.

**Figure 3.**
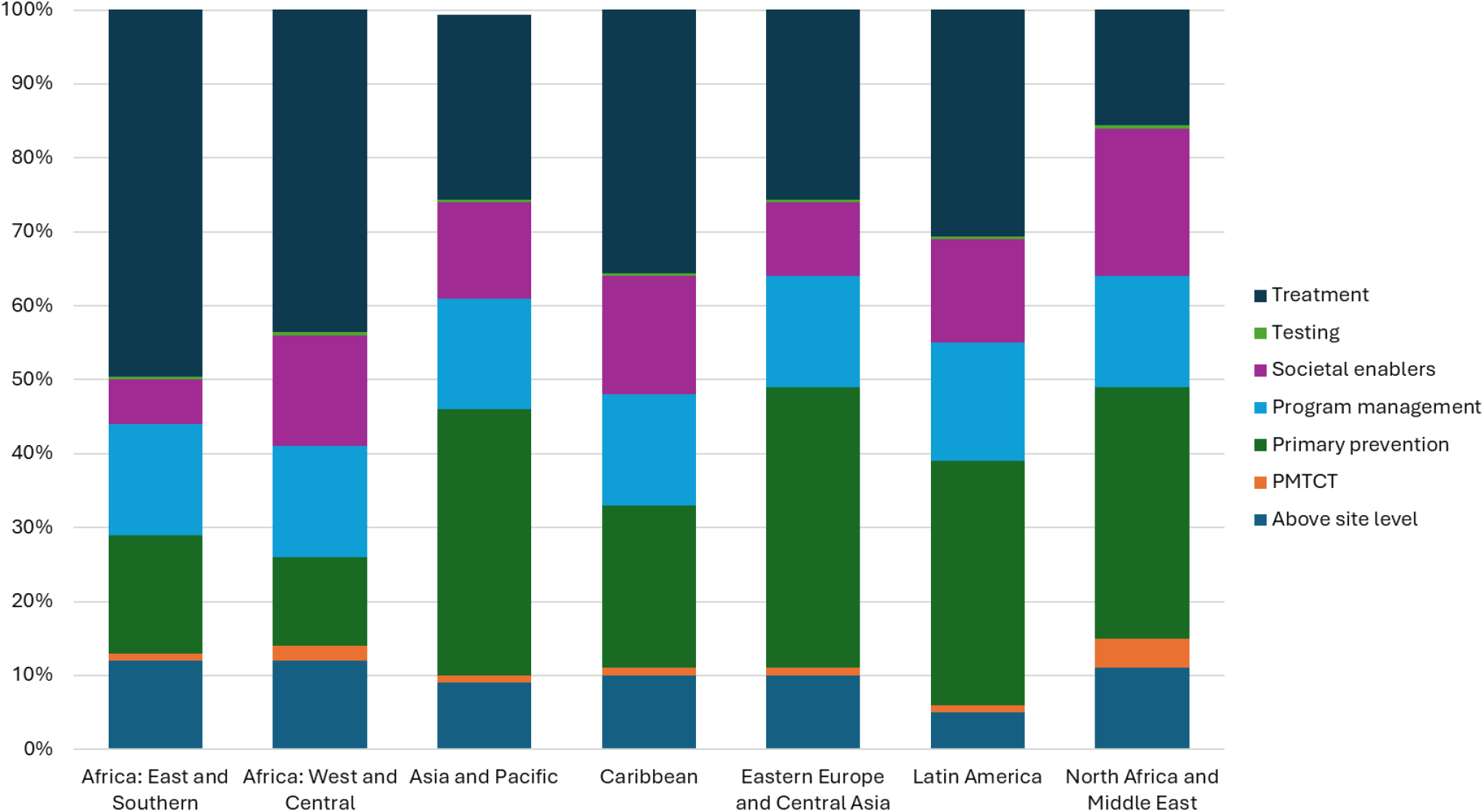
Percentage distribution of estimated resource needs in 2030 by region and intervention.

Commodities alone account for US$ 4.85 billion including US$2.8 billion for ARVs. (Table 3).

**Table 3.** Commodity costs in 2030 to meet the targets.

| Commodity | Cost in 2030 (Millions of US\$) |
| --- | --- |
| ARVs | \$2800 |
| Viral load tests | \$35 |
| CD4 tests | \$6 |
| Creatinine tests | \$38 |
| CrAg tests | \$3 |
| Rapid HIV tests | \$53 |
| Dual HIV-syphilis tests | \$179 |
| Early infant diagnosis tests | \$30 |
| Condoms | \$160 |
| Needles and syringes | \$185 |
| Opioid maintenance drugs | \$125 |
| Oral PrEP | \$258 |
| Injectable PrEP | \$890 |
| Total | \$4750 |

The largest needs are in East and South Africa (45%) followed by Asia/Pacific (29%), West and Central Africa (10%), Latin America and the Caribbean (10%), Eastern Europe and Central Asia (3%) and North Africa and Middle East (2%). By income group the largest needs are for Upper Middle-income countries (45%) followed by Lower Middle-income countries (39%) and Low-Income countries (16%).

Treatment remains the intervention with the largest share of resource needs in 2030 across most regions, accounting for 50% of total needs in Eastern and Southern Africa and 44% in Western and Central Africa, but a lower share of just 16% in the Middle East and North Africa. In contrast, primary prevention remains the major priority in Asia and the Pacific (36%), Eastern Europe and Central Asia (38%), Latin America (33%), and the Middle East and North Africa (34%). Societal enablers were particularly prominent in the Middle East and North Africa (20%), West and Central Africa (15%) and the Caribbean (16%).

## Discussion

Achieving the UNAIDS targets for 2030 would have substantial epidemiological impact and nearly achieve the goals of reducing new infections and AIDS deaths by 90% from 2010 to 2030. New infections would be just 9% of the peak value in 1995 and AIDS deaths would be 13% of the peak value in 2004. In 2030 new infections would be 60% less than deaths to PLHIV, setting us on a path to achieve the end of AIDS as a public health threat.

These estimates update previous work that estimated that new infections would fall to 370,000 new infections and 250,000 AIDS deaths by 2025 [2]. Although progress in recent years has not been as rapid as envisioned in 2020, we still have an opportunity to reach ambitious targets by 2030 if we act now.

The resources required to achieve these targets are substantial but $ 5 billion less than previous estimates of $ 29.3 billion. The main differences are due to price reductions observed for ARVs during the 2021-2024 period, lower unit costs for NSP and OAMT programs, a decision to limit condoms needs to the cost of commodities rather than including the distribution and promotion costs which are now contained in the health system costs, a geographical prioritization of key population intervention coverage based on HIV risk, and prioritization of societal enabler interventions in geographies based on unfavorable societal enabling index In this iteration of the resource needs estimates, Bulgaria and the Russian Federation which were reclassified as high-income by the World Bank in July 2024 were excluded from the analysis, which results in a lower overall projection of resource needs.

The detailed resource needs by intervention, population group, and country income level provide a foundation for countries to benchmark their current and projected domestic allocations. The resource needs estimates are critical not only for assessing the investment required to achieve the 2030 HIV targets, but also for gauging the feasibility of newly proposed domestic financing goals, based on the 2024 Global Task Force recommendation to increase domestic HIV spending, which will require careful alignment of available resource envelopes, projected needs, fiscal space, and national prioritization frameworks.

UNAIDS estimates that a total of $18.7 billion was available for HIV programs in low- and middle-income countries (LMICs) in 2024 [8]. That figure includes some donor administration expenditures that are not included in our estimates of in-country expenditures but it is clear that HIV resources need to increase by more than $3 billion annually by 2030 to meet these targets. Investments in HIV prevention must increase substantially to achieve the necessary scale required to achieve 2030 financing targets, rising to US$ 5.3 billion from the current level estimated by UNAIDS (US$ 1.7 to US$ 2.2 BN). Additionally, funding for societal enablers, currently estimated by UNAIDS as less than 5% of total HIV resources in 2024, needs to double, reaching 10% of the annual resource needs by 2030 to meet the financing targets on societal enablers. Many donors announced reductions to assistance for HIV programs in 2025. Thus, the task of mobilizing these additional resources is challenging, but if it can be done, we can achieve substantial success in combating this epidemic which has affected so many lives since it began 54 years ago.

## Data Availability

Data are summarized in the manuscript. The unit cost database is available on GitHub at https://github.com/JGStover/HIV-Unit-Costs/tree/main. Country-specific models are available upon request.

## References

1. https://aidsinfo.unaids.org/. Accessed June 30, 2025.

2. UNAIDS, Prevailing Against Pandemics, World AIDS Day Report, 2020: December 2020. Available from https://www.unaids.org/en/resources/documents/2020/prevailing-against-pandemics

3. Stover J, Glaubius R, Teng Y, Kelly S, Brown T, Hallett TB, et al. (2021) Modeling the epidemiological impact of the UNAIDS 2025 targets to end AIDS as a public health threat by 2030. PLoS Med 18(10): e1003831. 10.1371/journal.pmed.1003831.

4. UNFPA, Costing The Three Transformative Results, January 2020, https://www.unfpa.org/featured-publication/costing-three-transformative-results.

5. Remme M, Vassall A, Lutz B, Luna J, Watts C. Financing structural interventions: going beyond HV-only value for money assessments. AIDS 2014, 28:425–434.

6. World Health Organization. UHC Service Coverage Index. Updated 9 January 2024. Available at https://data.who.int/indicators/i/3805B1E/9A706FD.

7. United Nations, Department of Economic and Social Affairs, Population Division (2004). Available at https://population.un.org/wpp/

8. UNAIDS. Financial Dashboard. Accessed on June 30, 2025. Available at https://aidsinfo.unaids.org/finance

9. https://github.com/JGStover/HIV-Unit-Costs/tree/main

10. Global Task Team on 2030 HIV Targets Writing Group. The Lancet. Forthcoming.

11. Eaton JW, Dwyer-Lindgren L, Gutreuter S, et al. Naomi: a new modelling tool for estimating HIV epidemic indicators at the district level in sub-Saharan Africa. J Int AIDS Soc. 2021;24:25788.

