## Supplemental Text for "The impact and cost of reaching the UNAIDS global HIV targets"

**Scope of interventions, programmes and activities**

A scoping literature review was undertaken in Pubmed, Web of Science and Scopus to identify studies with proven impact on reducing HIV and KP related stigma and discrimination. The search strategy was based on the Population, Concept, Context framework recommended by the Joanna Briggs Institute for scoping reviews using related keywords and search terms covering the period until 1 December 2024. Additional studies were also included following consultation with HIV stigma experts.

We identified 17 social enabling interventions that were considered for costing and were summarized under five programme areas as follows:

- People living with HIV to reduce internalized stigma (Six programmes [1-6], Table S1)
- People living with HIV to reduce experienced stigma and discrimination in healthcare and community settings (Two programmes [7, 8], Table S2)
- KPs to reduce experienced stigma and discrimination (Two programmes [7, 9] , Table S3)
- Reduce discriminatory attitudes towards people living with HIV in the general population (Two programmes [10, 11] , Table S4)
- Reduce negative attitudes towards people living with HIV and key populations among health workers (Six programmes [12-17], two of those for healthcare students in tertiary education [16, 17] , Table S5)

One study [7] was considered for both PLHIV and KPs. These studies were focused on four population groups: KPs (men who have sex with men (MSM), female sex workers (FSWs), transgender people (TG)) [7, 9], PLHIV [1-8], healthcare students [16, 17] and healthcare/medical staff [12-15], and the general population [10, 11]. For KPs we assumed that these RNEs also represent people who inject drugs (PWID) and prisoners, although these two groups were not included in the 17 interventions that were costed. Only HIV and stigma & discrimination related activities were costed in the selected interventions, which were costed per population group and HIV stigma domain to avoid double costings, e.g. start up intervention costs, and to ensure that interventions were targeted to the appropriate population group.

**Assumptions for estimating costs in the 15 GF countries**

| **Assumption** | **Details** |
| --- | --- |
| Unit costs | Unit costs from the list of 15 GF countries are similar to unit costs in other countries. Variations in unit costs between countries were taken into consideration by estimating the average cost of interventions across countries by income level. |
| Cost components | Three categories of costs:  Start-up costs (eg. development of a curriculum)  Recurrent costs ((eg. media campaigns)  Variable costs (eg. costs for x number of workshops needed to reach between 40% to 80% of the target population by 2031)  Attributing scaling up costs may vary considerably by country and by intervention. An average intervention cost was assumed for each of the three cost categories across the 15 GF countries |
| Outputs | Information on outputs (eg. number of PLHIV reached) was extracted from the identified studies and it was assumed that interventions can be delivered in all 15 GF countries under the same circumstances irrespective of the context. |
| Context | Location or context was not considered when extrapolating unit costs from the 15 GF countries to the 17 social enabling interventions |

**Assumptions on intensity and scale up to reach targets in KPs to 2031**

The composite Societal Enabler Index was used to identify whether a country has a less or more favourable societal enabling environment. Countries were assigned into one of four groups based on quartiles of the composite Societal Enabler Index, with the most intense scale-up taking place in countries with less favourable societal enabling environment. Coverage was scaled up progressively from 40% in 2026 to 80% in 2031, with countries in the most intense scale up group reaching 80% by 2028. An increase of 40% coverage for KPs from 2026 to 2031 was considered assuming that each individual will attend a programme at least once every 2.5 years (i.e. 40% coverage annually). For healthcare workers, it was assumed that one of the interventions would be delivered annually and the number of hospitals to be implemented was based on country’s population density to account for vast territories with scattered communities. For healthcare students, one intervention would be delivered to tertiary education institutions and the number of institutions was based on country’s population density to account for vast territories with scattered communities. For general population, interventions were delivered annually assuming that they cover over 90% of the general population (ie. mass media campaigns).

Countries selected to implement the least expensive of the interventions that were costed in each programme area, to account for the current resource constrained environment, except the two interventions for the general population that are combined together to produce one cost.

| **Scale up intensity** | **2026 (baseline coverage)** | **2027** | **2028** | **2029** | **2030** | **2031** |
| --- | --- | --- | --- | --- | --- | --- |
| **Least intense** | 0.4 | 0.5 | 0.6 | 0.7 | 0.8 | 0.8 |
| **2** | 0.4 | 0.5 | 0.7 | 0.7 | 0.8 | 0.8 |
| **3** | 0.4 | 0.6 | 0.7 | 0.8 | 0.8 | 0.8 |
| **Most intense** | 0.4 | 0.6 | 0.8 | 0.8 | 0.8 | 0.8 |

| **Most intense (4)** | **3** | **2** | **Least intense (1)** |
| --- | --- | --- | --- |
| Afghanistan | Albania | Armenia | Argentina |
| Benin | Algeria | Azerbaijan | Belize |
| Burkina Faso | Angola | Bangladesh | Botswana |
| Central African Republic | Burundi | Belarus | Brazil |
| Chad | Cabo Verde | Bhutan | Bulgaria |
| Congo, Dem. Rep | Cameroon | Bolivia | Cambodia |
| Congo, Rep. | Egypt, Arab Rep. | Bosnia and Herzegovina | China |
| Côte d'Ivoire | Fiji | Ecuador | Colombia |
| Equatorial Guinea | Gambia, The | Honduras | Comoros |
| Eritrea | Guatemala | Iran, Islamic Rep. | Costa Rica |
| Eswatini | Indonesia | Kazakhstan | Cuba |
| Ethiopia | Jamaica | Korea, Dem. People's Rep. | Dominican Republic |
| Gabon | Jordan | Kyrgyz Republic | El Salvador |
| Ghana | Lao PDR | Lebanon | Georgia |
| Guinea | Malawi | Lesotho | Guyana |
| Guinea-Bissau | Moldova | Libya | India |
| Haiti | Mongolia | Nepal | Kenya |
| Iraq | Morocco | Peru | Malaysia |
| Liberia | Mozambique | Rwanda | Maldives |
| Madagascar | Myanmar | Serbia | Mexico |
| Mali | Nigeria | Suriname | Montenegro |
| Mauritania | North Macedonia | Tunisia | Namibia |
| Niger | Pakistan | Turkey | Nicaragua |
| Papua New Guinea | Philippines | Ukraine | Paraguay |
| Senegal | Syrian Arab Republic | Uzbekistan | Russian Federation |
| Sierra Leone | Tajikistan | Zimbabwe | South Africa |
| Somalia | Tanzania |  | Sri Lanka |
| South Sudan | Turkmenistan |  | Thailand |
| Sudan | Uganda |  | Venezuela, RB |
| Timor-Leste | Zambia |  | Vietnam |
| Togo |  |  |  |
| Yemen, Rep. |  |  |  |

**Assumptions for producing global RNEs to 2031**

| **Assumption** | **Value** |
| --- | --- |
| Coverage of KPs at baseline | 40% |
| Intensity | Quintiles of the composite social enabler index |
| Estimated costs | Using the least expensive programme per programme area (except for programmes in the general population) |
| DL training/workshops | Internet use below median (<62.6%), delivered up to five times the number of people compared to F2F  Internet use above median (>62.5%), delivered up to ten times the number of people compared to F2F |
| Number of tertiary education institutions to deliver programmes for healthcare students | Countries with population density above 101 people per sq km: 2 institutions  Countries with population density between 47 and 100 people per sq km: 3 institutions  Countries with population density below 47 people per sq km: 5 institutions |
| Number of primary schools to deliver the programmes for the general population | Countries with population density above 101 people per sq km: 30 primary schools  Countries with population density between 47 and 100 people per sq km: 50 primary schools  Countries with population density below 47 people per sq km: 100 primary schools |
| Number of hospitals delivering ART services | Countries with population density above 101 people per sq km: 5 hospitals  Countries with population density between 47 and 100 people per sq km: 20 hospitals  Countries with population density below 47 people per sq km: 50 hospitals |
| N of mass media campaigns | Countries with population density above 101 people per sq km: 1 campaign  Countries with population density between 47 and 100 people per sq km: 3 campaigns  Countries with population density below 47 people per sq km: 5 campaigns |
| N of NGOs delivering ART services | HIV prevalence < 1% (Low): 1  HIV prevalence between 1 and 2% (Moderate): 3  HIV prevalence over 2% (High): 5 |
| Cost adjustments based on HIV prevalence* | HIV prevalence < 1% (Low): 25%  HIV prevalence between 1 and 2% (Moderate): 50%  HIV prevalence over 2% (High): 100% |

**Applied to the following programme areas:*

*For people living with HIV to reduce internalized stigma (n = 6)*

*For people living with HIV to reduce experienced stigma and discrimination in healthcare and community settings (n = 2)*

*For KPs to reduce experienced stigma and discrimination (n = 2)*

**Distribution of intervention costs by programme area**

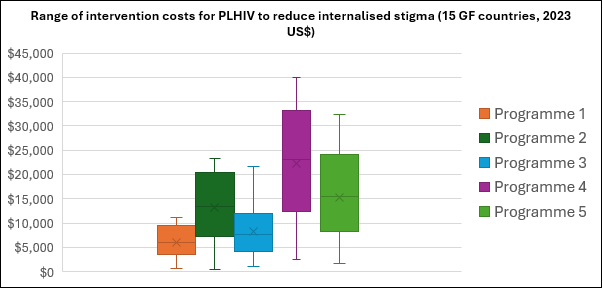

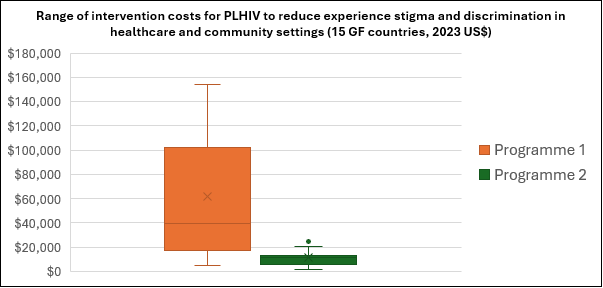

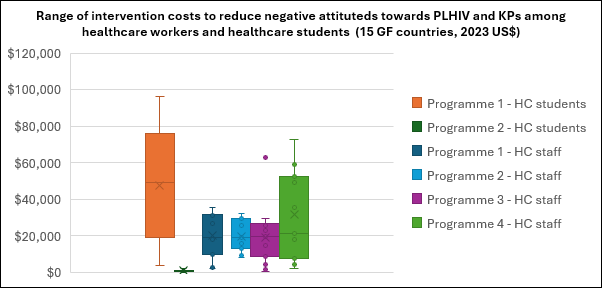

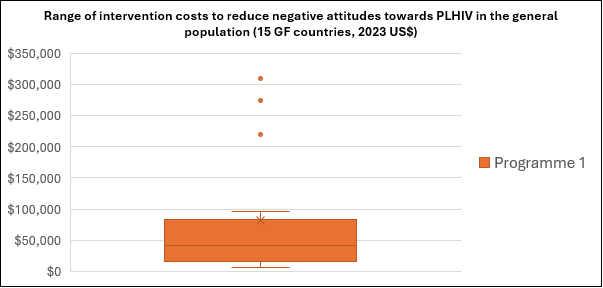

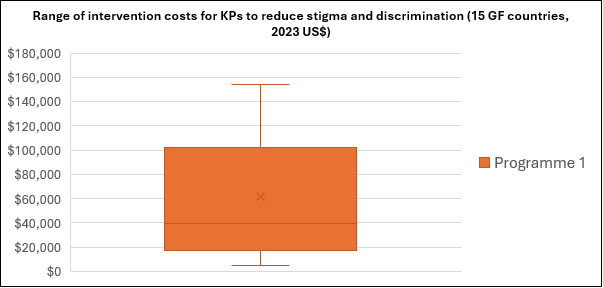

Resource Needs Estimates 2026-2031

Millisons (2023 US$)

|  | Programmes for people living with HIV to reduce internalised stigma | Programmes for people living with HIV to reduce experienced stigma and discrimination in healthcare and community settings | Programmes for key populations to reduce experienced stigma and discrimination | Programmes to reduce discriminatory attitudes towards people living with HIV in the general population | Programmes to reduce negative attitudes towards people living with HIV and key populations among health workers |
| --- | --- | --- | --- | --- | --- |
| 2026 | 1.38829 M | 10.8487 M | 7.07126 M | 153.431 M | 89.7594 M |
| 2027 | 0.949531 M | 11.0207 M | 1.12492 M | 151.89 M | 90.2444 M |
| 2028 | 0.972755 M | 12.1321 M | 1.13876 M | 153.712 M | 91.3273 M |
| 2029 | 0.988257 M | 12.9235 M | 0.521763 M | 155.557 M | 92.4233 M |
| 2030 | 1.0051 M | 13.452 M | 0.528109 M | 157.423 M | 93.5324 M |
| 2031 | 1.01838 M | 13.647 M | 0.534533 M | 159.313 M | 94.6547 M |

Table S1. Programmes to reduce internalised stigma in people living with HIV

| **Author** | **Programme description** | **Cost components** | **Cost category** | **Unit cost source, inputs and outputs** | **Comments and assumptions** |
| --- | --- | --- | --- | --- | --- |
| Barroso et al, 2013 ^1^  Programme 1 | A 45-min video titled, ‘‘Maybe Someday: Voices of HIV-Positive Women.’’ It portrays five composite representations of HIV-Positive women with each relating a narrative based on one or two themes derived from the synthesized studies. Characters in the video are designed to connect with viewers on multiple levels and acknowledge the interplay, connections, and potential disconnections between their HIV status and other aspects of their lives. Each character shares difficult personal details with an off-camera listener and affords viewers the privilege of witnessing her reflections and, in some cases, decision making. | Research survey. Upon return of the surveys, participants were given a $10 gift card to thank them for their time. | Start up cost | Source: Barroso et al, 2013  Outputs: 99 women | Research costs related to the survey to help design and develop the video, which is a start up one off cost in Year 1. It is assumed that costs for gift cards and iPods are similar in countries and these costs are part of a survey run in the first year.  The 45 min video was assumed to be a cost related to media campaign through radio/IEC.  Every year a community awareness campaign to disseminate the video will be carried out, assuming that the campaign covers 90% of the women living with HIV who experience internalized stigma |
|  |  | Distributing flyers at a tertiary care infectious diseases clinic to recruit 10–15 HIV-infected women (who met the inclusion criteria for the primary study) to participate in a pre-intervention focus group, to obtain data for refining study instruments and procedures. Women who participated in the focus group received a $25 gift card | Start up cost | Source: Barroso et al, 2013  Outputs: 99 women |  |
|  |  | Kept the iPod as a thank you for participating in the study | Start up cost | Source: Barroso et al, 2013  Outputs: 99 women |  |
|  |  | Develop a 45-min video | Start up cost | Source: Barroso et al, 2013 |  |
|  |  | Conduct community awareness campaign/road show including IEC materials (ie. video) | Recurrent annual cost | Source: Global Fund |  |
| Bhatta & Liabsuetrakul, 2016 ^2^  Programme 2 | Empowerment intervention social self-value package with an aim to improve the quality of life (QoL) of HIV infected people receiving antiretroviral treatment.  The empowerment intervention mainly focused on autonomy and community activism, self-esteem/self-efficacy, self-care, optimism and control over the future, family and social relationships, power-powerlessness, management of stress and righteous anger, stigma and discrimination issues, legal provisions, and human and health rights.  Participants receive six weekly intervention sessions. The intervention is delivered over six sessions held weekly at the ART center lasting one and half hours. Sessions are conducted with a group of 8–10 participants. A debriefing session is conducted at the end of each session for the feedback from the reviewer and facilitators. A facilitator delivers the intervention with participatory learning activities, buzz sessions, brain storming, lecture, and discussion techniques. Participants are encouraged and motivated to communicate and discuss with different people about prevention, treatment and disclosure of HIV issues. | All the sessions were facilitated by two national level trainers with a public health graduate degree. | Cost to scale up | Source: Global Fund  Inputs: Two facilitators  Outputs: 66 PLHIV receiving ART | Debriefing is undertaken in all sessions in Year 1. Feedback was used to enhance the content of the sessions, which is covered in the overall cost. |
|  |  | A debriefing session was conducted at the end of each session for the feedback from the reviewer and facilitators. | Start up cost | Source: Global Fund  Inputs: Two facilitators  Outputs: 66 PLHIV receiving ART |  |
| Chidrawi et al, 2016 ^3^  Programme 3 | The intervention is adapted from the validated intervention manual of Uys et al. (2009) and is based on three tenets namely the a) sharing of information on HIV stigma and coping with HIV stigma, b) the equalising of relationships between PLWH and PLC through increased interaction and contact among them by grouping them together, and c) the empowerment of members of both groups towards leadership in HIV stigma reduction through practical knowledge and experience of project planning regarding HIV stigma reduction and implementation in their communities. | A two-day presentation and activity-based workshop for PLHIV only. It focused on their personal understanding of HIV stigma, identification of their personal strengths and teaching responsible disclosure management to prepare them for the rest of the workshops in the intervention | Start up cost | Source: Global Fund  Inputs: Two facilitators  Outputs: 60 PLHIV | The two-day presentation and activity-based workshop was delivered by two facilitators to 60 people living with HIV or AIDS in Year 1. |
|  |  | Six three-day workshops for each group of people living close (PLC) to people living with HIV or AIDS. The PLC workshops occurred two weeks apart and were led by two facilitators (one HIV-infected and one non-HIV infected person) for each group. These workshops were attended by all PLHIV. | Cost to scale up | Source: Global Fund  Inputs: Two facilitators  Outputs: 60 PLHIV |  |
|  |  | In the third one-day workshop, the original designated group invited community members as guests and then presented feedback on their projects. | Cost to scale up | Source: Global Fund  Inputs: Two facilitators  Outputs: 60 PLHIV |  |
| Ferris France et al, 2019 ^4^  Programme 4 | Intervention, consisting of a 12-week facilitated programme using a guided form of self-inquiry which helps users to overcome negative thoughts and beliefs such as Self-stigma–negative self-judgements resulting in shame, worthlessness and self-blame– may play a crucial role in emotional reactions and cause emotional distress | Two local facilitators worked with international certified facilitators to deliver and adapt the programme with two groups of participants (23 in total) drawn from support groups. | Start up cost | Source: Global Fund  Inputs: Two facilitators  Outputs: 23 people | A workshop to develop the intervention manual and train facilitators to deliver the workshops. Costs incurred in Year 1. |
|  |  | The participants took part in 12 four hour group sessions run over 12 weeks | Cost to scale up | Source: Global Fund  Inputs: Two facilitators  Outputs: 23 people |  |
|  |  | A weekly one-hour individual session with a facilitator, as well as homework. | Cost to scale up | Source: Global Fund  Inputs: Two facilitators  Outputs: 23 people |  |
| Rao et al, 2012 ^5^  Programme 5 | Adapted intervention to reduce internalized stigma for women living with HIV.  A workshop that met for 4–5 h during 2 consecutive weekday afternoons. Participants are scheduled to participate in 1 of 2 workshops. | Adapting the intervention in Year 1 | Start up cost | Source: Global Fund  Inputs: 90 people | A workshop to develop the intervention manual and train facilitators to deliver the workshops. A total of 30 peer educations and 60 social workers were trained. Costs incurred in Year 1. The first and second workshops are delivered to women living with HIV by one peer facilitator and one social worker. |
|  |  | First 4-5 hr workshop | Cost to scale up | Source: Global Fund  Inputs: Two facilitators Outputs: 13 women living with HIV |  |
|  |  | Second 4-5 hr workshop | Cost to scale up | Source: Global Fund  Inputs: Two facilitators Outputs: 13 women living with HIV |  |
| Smith Fawzi et al, 2012 ^6^  Programme 6 | A psychosocial support group intervention for HIV-affected youth and their caregivers taking place in a NGO delivering ART services. | Phase 1 consists of eight group sessions of HIV-positive caregivers, focusing on healthy lifestyles, coping with negative feelings, disclosing their HIV status, along with other challenges of being HIV-positive. | Cost to scale up | Source: Global Fund  Inputs: Three facilitators Outputs: 298 people | The number of facilitators is not given in the programme, and it is assumed that one psychologist and two social workers are needed to run the sessions. It is assumed that a conference room is needed per day for the 10 groups of 15 pairs (parent-child) for each of the three sessions (one session in phase 1 and two sessions in phase 2). |
|  |  | Children during the first part of Phase 2 meet for seven sessions and focus on strategies for reducing emotional stress, developing a broader range of coping strategies, and reducing HIV risk behavior. During the second part of Phase 2, children and their caregivers meet together for eight sessions and focus on parent-child communication, conflict resolution, as well as prevention of risk behaviors related to early pregnancy, transmission of HIV and other STDs, as well as drug and alcohol abuse. | Cost to scale up | Source: Global Fund  Inputs: Three facilitators Outputs: 298 people |  |

Table S2. Programmes for people living with HIV to reduce experienced stigma and discrimination in healthcare and community settings

| **Author** | **Programme description** | **Cost components** | **Cost category** | **Unit cost source, inputs and outputs** | **Comments and assumptions** |
| --- | --- | --- | --- | --- | --- |
| Adam et al, 2011 ^7^  Programme 1 | Create a community mobilisation model to engage the discourses of moral reasoning and sexual decision making circulating in local communities of gay and bisexual men and to stimulate community building by providing a forum for dialogue that could affect local cultures to enhance sexual health.  The primary forum for the generation of the intervention is a series of meetings of a broad-based consortium of community members drawn from frontline HIV prevention work, along with representatives from public health, government and research plus staff support from the provincial government, the MSM community and a marketing design firm. | Prevention workers from partner organisations and the health ministry including sexual health clinics, public health units and other community-based organizations with HIV programming or gay clientele, participate in a one-day orientation workshop in advance of the campaign launch | Start up cost | Source: Global Fund  Outputs: 25 people | There is one off start up cost to inform the media advertising and website development. The HIV campaign will run over 10 years, with website and advertising content updated and costs incurred every 3-4 years. All components of the campaign, except community outreach, are not scaled up as it is assumed that the website reaches out to all MSM.  There is provision for targeted internet-based information, education, communication (social media groups, 24 hour helpline). Maintaining website over 10 years, including annual costs for moderators and blog facilitators. |
|  |  | Traditional media advertising (graphic is circulated on billboards, in print media (gay, entertainment and ethnocultural media in multiple languages), in online advertisements and in outreach materials) | Start up cost | Source: Global Fund |  |
|  |  | Community outreach (HIV prevention workers distribute materials to gay venues across the province) | Cost to scale up | Source: Global Fund  Inputs: 25 people  Outputs: 1942 MSM |  |
|  |  | Website (development of an attractive, professional innovative web-supported stigma reduction intervention and interactive website that would provide essential information, referrals and a community forum) | Recurrent annual cost | Source: Global Fund |  |
| Hosek et al, 2011 ^8^  Programme 2 | The intervention Project ACCEPT (Adolescents Coping, Connecting, Empowering, and Protecting Together), encompasses primary, secondary, and tertiary prevention among HIV-infected and HIV at-risk preadolescents, adolescents, and young adults up to 25 years of age.  The ACCEPT Curriculum included orientation sessions and sessions on cohesion & HIV overview, disclosure, preparing for medical intervention, healthy living, stress, relaxation and spirituality, female sexuality, male sexuality, self-esteem & self-worth, legal aid & advocacy, and future planning. | Each participant completes the first individual session. | Cost to scale up | Source: Global Fund  Inputs: Two facilitators  Outputs: 50 young adults | It assumed that the programme is delivered by two facilitators. |
|  |  | The study coordinator then schedules a time for the participant to complete the second individual session. | Cost to scale up | Source: Global Fund  Inputs: Two facilitators  Outputs: 50 young adults |  |
|  |  | After the second individual session is completed, the participant joins the group and participate in nine sessions. | Cost to scale up | Source: Global Fund  Inputs: Two facilitators  Outputs: 50 young adults |  |
|  |  | After the group sessions concluded, each participant is scheduled for the third individual session. | Cost to scale up | Source: Global Fund  Inputs: Two facilitators  Outputs: 50 young adults |  |

Table S3. Programmes for key populations to reduce experienced stigma and discrimination

| **Author** | **Programme description** | **Cost components** | **Cost category** | **Unit cost source, inputs and outputs** | **Comments and assumptions** |
| --- | --- | --- | --- | --- | --- |
| Adam et al, 2011 ^7^  Programme 1 | *See above* | *See above* | *See above* | *See above* | *See above* |
| Catalani et al, 2012 ^9^  Programme 2 | This programme is delivered in three phases: the development of a feature film Prarambha (The Beginning), the development of an illustrated stigma video, and the subsequent pilot testing of both media. Two HIV stigma videos are created using techniques from traditional film production and new media digital storytelling. A series of 16 focus group discussions are held, with specific groups for sex workers, men who have sex with men, young married women, and others. Focus groups with viewers of the traditional film (8 focus groups, 80 participants) and viewers of the new media production (8 focus groups, 69 participants) revealed the mechanisms through which storyline, characters, and aesthetics influence viewers’ attitudes and beliefs about stigma. | This illustrated video is developed during four production stages by a team of health communication experts, a local artist, different community translators representing four local languages, and voice over artists to depict female and male voices in local languages. Pre-production includes a myriad of activities from fund raising and contract negotiation, to recruitment of contributors and coordination of their schedules, to storyline development and screenwriting. The film was supported by an international organisation for under $250,000. | Start up cost | Source: Paper | Research costs related to the survey to help design and develop the film and illustrated video, which is a start up one off cost in Year 1.  It is assumed that a similar budget will be required to develop and produce the film and video, including staff time, in different countries.  Every year a community awareness campaign to disseminate the film and video will be carried out, assuming that the campaign covers 90% of KPs. |
|  |  | Based on recommendations from local partners, participants were not remunerated, however they were served a lunch and provided with a gift valuing $2–4 USD. | Start up cost | Source: Paper |  |
|  |  | Production, which begins with an established story line, take approximately 2 months and cost $8,000, including staff time. | Start up cost | Source: Paper |  |
|  |  | Conduct community awareness campaign/road show including IEC materials (ie. film and video) | Recurrent annual cost | Source: Global Fund |  |

Table S4. Programmes to reduce discriminatory attitudes towards people living with HIV in the general population

| **Author** | **Programme description** | **Cost components** | **Cost category** | **Unit cost source, inputs and outputs** | **Comments and assumptions** |
| --- | --- | --- | --- | --- | --- |
| Bell et al, 2007 ^10^  Programme 1 | The CHAMPSA intervention targeted HIV risk behaviors by strengthening family relationship processes as well as targeting peer influences through enhancing social problem solving and peer negotiation skills for youths | Each school received a school improvement stipend of $1,000. | Start up cost | Source: Paper | Rewards were provided assuming that all caregivers attended all 10 sessions. It is assumed that stipends, rewards and incentives have similar cost in different countries. |
|  |  | The final adapted CHAMPSA manualized program, comprises 10-90-minute sessions delivered over 10 weekends. Given the amount of time families are required to devote to the program, they were paid a stipend of $8 for each session attended | Cost to scale up | Source: Paper  Output: 478 primary school students (20 primary schools) |  |
|  |  | $30 incentive for attending all 10 sessions. | Cost to scale up | Source: Paper  Output: 478 primary school students (20 primary schools) |  |
| Fakolade et al, 2010 ^11^  Programme 2 | Mass media involving the dissemination of key preventive messages, the active involvement of faith-based organizations (FBOs), workplace interventions and private sector participation. ‘Exposure to mass media messages on HIV and AIDS’ based on viewer-ship, listenership and intensity (frequency) of being exposed to all or some of the HIV and AIDS messages aired. | Mass media campaigns through radio and other IEC material. | Recurrent annual cost | Source: Global Fund | All components of the mass media campaign are not scaled up as it is assumed that mass media reaches out to everyone. |

Table S5. Programmes to reduce negative attitudes towards people living with HIV and key populations among healthcare workers and students

| **Author** | **Programme description** | **Cost components** | **Cost category** | **Unit cost source, inputs and outputs** | **Comments and assumptions** |
| --- | --- | --- | --- | --- | --- |
| Ezedinachi et al, 2002 ^12^  *Healthcare workers*  Programme 1 | Intervention and dissemination of training in clinical management, health education, and attitudinal change toward patients with HIV disease. | Following initial questionnaire-defining focus groups, nurses, laboratory technologists and physicians in hospitals are trained by influential role models who attended the initial training. Two main ‘Train the Trainer’ workshops are held in the capital and regional cities. | Start up cost | Source: Global Fund  Input: Two facilitators  Output: 1072 medical staff (One high level hospital) | The initial two-day train the trainer workshop is delivered assuming two trainers/facilitators are involved in delivering this per local governmental area. This was costed in Year 1. Two workshops are delivered to 20 participants. Two participants then deliver two smaller workshops in each of the local areas reaching 1072 health workers. This training is assumed to take place in one high-level urban hospital. |
|  |  | Smaller workshops and seminars, two in different admin areas, are delivered by those trained in the initial training. | Cost to scale up | Source: Global Fund  Input: Two facilitators  Output: 1072 medical staff (One high level hospital) |  |
| Nyblade et al, 2020 ^13^  *Healthcare workers*  Programme 2 | Targeting the whole facility beyond HIV services, the intervention approach includes a two-day participatory stigma-reduction training for all staff levels (clinical and non-clinical) with delivery by staff and clients from the facilities who are trained as stigma-reduction facilitators. Trainings are delivered to all categories of staff, with a target of reaching 70% of the facility workforce. The trainings are based on pre-existing global training materials that are available online | In-country stakeholder workshop to adapt intervention manual. | Start up cost | Source: Global Fund  Output: Two facilitators and 20 representatives of the health ministry and other organisations | The 1-day stakeholder workshop to adapt and develop the intervention manual to deliver at the national level takes place in Year 1. It is assumed that the workshop was facilitated by two stigma experts and 20 reps from different stakeholders. Participatory data dissemination and review workshops with staff is undertaken in the hospital. It is assumed that these are 1 day workshop facilitated by two people and running once every year in the hospital. Additionally, a two-day participatory stigma-reduction training for all staff levels is carried out in the hospital every year and is facilitated by two people. The workshop are run in hospitals. Each facility is provided $5000 USD to develop facility-specific ancillary activities. |
|  |  | Participatory data dissemination and review workshops with staff were undertaken in each hospital. It was assumed that these are 1 day workshop facilitated by two people and running once every year in each hospital. Additionally, a two-day participatory stigma-reduction training for all staff levels is carried out in every hospital every year and is facilitated by two people. | Cost to scale up | Source: Global Fund  Input: Two facilitators  Output: 243 medical staff (One high level urban hospital) |  |
|  |  | Each facility also created an eight to ten member “champion team,” which was provided $5000 USD to develop facility-specific ancillary activities, including launch events, anti-stigma-and-discrimination banners and posters, additional staff trainings, printed codes of ethics, reporting mechanisms, and staff nametags to enable identification and reporting of stigma and discrimination. | Cost to scale up | Source: Paper  Input: Two facilitators  Output: 243 medical staff (One high level urban hospital) |  |
| Pulerwitz et al, 2015 ^14^  *Healthcare workers*  Programme 3 | Intervention include staff training (ranging from physicians to ward cleaners), hospital policy development, and supplies provision focusing on ‘fear-based’ stigma (stemming from lack of knowledge) and social stigma (stemming from moral judgments). | Hospital staff receive a half-day training on HIV transmission and prevention, and a full day on appropriate infection control measures. Hospital staff also participate in an extra half-day training on social stigma co-facilitated by persons living with HIV (e.g., more emphasis on the perspectives of PLHIV patients). There are three main categories of staff trainees: doctors and nurses, auxiliary nurses/ward staff, and administration and support staff. Each training group ranged in size from 25 to 45 people, depending on the number of staff, and from 6 to 10 training groups were organized in each facility. | Cost to scale up | Source: Global Fund  Input: Two facilitators  Output: 25 to 45 medical staff (One high level urban hospital) | A total of two days workshop in each hospital every year. Each training group ranged in size from 25 to 45 people, depending on the number of staff, and from 6 to 10 training groups were organized in each facility. It is assumed that on average there were eight 2d workshops per hospital per year delivered by two facilitators/trainers.  The policy document and processes are reviewed every year. |
|  |  | Hospital policy development document in line with the AIDS law (recently released at the time). Overall policy to focus on six policy components: (1) Access to services by people living with HIV, (2) HIV counseling and testing, (3) Confidentiality, (4) Appropriate infection control, (5) Training on HIV and AIDS, and (6) Dissemination of the HIV and AIDS-related policy. | Recurrent annual cost | Source: Global Fund  Input: Two facilitators |  |
| Uys et al, 2009 ^15^  *Healthcare workers*  Programme 4 | The intervention consists of bringing together a team of approximately 10 nurses and 10 people living with HIV or AIDS (PLHA) in each setting and facilitating a process in which they plan and implement a stigma reduction intervention, involving both information giving and empowerment.  The project initiation workshop is facilitated by a nurse and PLHA based on a standard manual and both are trained over a period of 2 days prior to them running the workshops. On the basis of this training the teams are then given the task of designing, implementing, and evaluating a project to reduce stigma in their health care setting within a month, with the support of the facilitators. The project concludes with a 1-day project evaluation workshop again facilitated by the facilitators. | Training costs 2-day workshop | Recurrent annual cost | Source: Global Fund  Input: Two facilitators  Output: 20 people, 10 nurses and 10 PLHIV (One high level urban hospital) | A nurse and a person living with HIV or AIDS were trained over 2 days to deliver the programme. Each workshop takes place in the hospital setting and is delivered to an average of 40 medical staff per hospital. |
|  |  | Training costs 1-day additional workshop | Cost to scale up | Source: Global Fund  Input: Two facilitators  Output: 40 medical staff (One high level urban hospital) |  |
| Arora et al, 2014 ^16^  *Healthcare students*  Programme 1 | Five day empowering programme  prepared in consultation with eight experts from community medicine and nursing field with an objective to expand the understanding of student nurses and modify their beliefs related to HIV/AIDS. First two days focused on the magnitude, basic dynamics, mode of transmission and prevention of HIV/AIDS. Next two days were dedicated to alter the beliefs of student nurses about HIV infection and AIDS. Lecture, group discussion and role play were used to impact correct information to the students about AIDS. The study was conducted among 33 students nurses pursuing third year BSc nursing and General nursing. | Development of programme manual | Start up cost | Source: Global Fund  Input: Eight experts | The programme is adapted during a one day workshop with eight experts in Year 1. The programme is delivered to a class of 33 healthcare students by one facilitator over 5 days. |
|  |  | Five day empowering programme was prepared in consultation with eight experts from community medicine and nursing field | Cost to scale up | Source: Global Fund  Input: Two facilitators  Output: 33 healthcare students (One tertiary education institute) |  |
| Mak et al, 2018 ^17^  *Healthcare students*  Programme 2 | A 30-minute didactic session on HIV/AIDS knowledge, followed by two different experiential games and a 90-minute sharing session hosted by two PLHIV. | Development of programme manual | Start up cost | Source: Global Fund  Input: Two experts | The programme is reviewed and adapted during a one day workshop with two experts in Year 1. Two PLHIV delivered the 90-minute sharing session to a class of healthcare students. The 30-minute didactic session on HIV/AIDS knowledge, and the two different experiential games were delivered to a class of healthcare students by one facilitator. |
|  |  | Two PLHIV hosting a 90-minute sharing session by a local NGO dedicated to improving the living standards of PLHIV | Cost to scale up | Source: Global Fund  Input: Two facilitator  Output: 46 healthcare students (One tertiary education institute) |  |
|  |  | A 30-minute didactic session on HIV/AIDS knowledge, followed by two different experiential games | Cost to scale up | Source: Global Fund  Input: One facilitators  Output: 46 healthcare students (One tertiary education institute) |  |

**References**

1.	Barroso J, Relf MV, Williams MS, et al. A randomized controlled trial of the efficacy of a stigma reduction intervention for HIV-infected women in the Deep South. *AIDS Patient Care STDS*. Sep 2014;28(9):489-98. doi:10.1089/apc.2014.0014

2.	Bhatta DN, Liabsuetrakul T. Efficacy of a Social Self-Value Empowerment Intervention to Improve Quality of Life of HIV Infected People Receiving Antiretroviral Treatment in Nepal: A Randomized Controlled Trial. *AIDS Behav*. Jun 2017;21(6):1620-1631. doi:10.1007/s10461-016-1546-z

3.	Chidrawi HC, Greeff M, Temane QM, Doak CM. HIV stigma experiences and stigmatisation before and after an intervention. *Health SA Gesondheid*. 2016;21:196-205. doi:10.1016/j.hsag.2015.11.006

4.	Ferris France N, Macdonald SH, Conroy RR, et al. 'We are the change' - An innovative community-based response to address self-stigma: A pilot study focusing on people living with HIV in Zimbabwe. *PLoS One*. 2019;14(2):e0210152. doi:10.1371/journal.pone.0210152

5.	Rao D, Desmond M, Andrasik M, et al. Feasibility, acceptability, and preliminary efficacy of the unity workshop: an internalized stigma reduction intervention for African American women living with HIV. *AIDS Patient Care STDS*. Oct 2012;26(10):614-20. doi:10.1089/apc.2012.0106

6.	Smith Fawzi MC, Eustache E, Oswald C, et al. Psychosocial support intervention for HIV-affected families in Haiti: implications for programs and policies for orphans and vulnerable children. *Soc Sci Med*. May 2012;74(10):1494-503. doi:10.1016/j.socscimed.2012.01.022

7.	Adam BD, Murray J, Ross S, Oliver J, Lincoln SG, Rynard V. hivstigma.com, an innovative web-supported stigma reduction intervention for gay and bisexual men. *Health Educ Res*. Oct 2011;26(5):795-807. doi:10.1093/her/cyq078

8.	Hosek SG, Lemos D, Harper GW, Telander K. Evaluating the acceptability and feasibility of Project ACCEPT: an intervention for youth newly diagnosed with HIV. *AIDS Educ Prev*. Apr 2011;23(2):128-44. doi:10.1521/aeap.2011.23.2.128

9.	Catalani C, Castaneda D, Spielberg F. Development and Assessment of Traditional and Innovative Media to Reduce Individual HIV/AIDS-Related Stigma Attitudes and Beliefs in India. *Front Public Health*. 2013;1:21. doi:10.3389/fpubh.2013.00021

10.	Bell CC, Bhana A, Petersen I, et al. Building protective factors to offset sexually risky behaviors among black youths: a randomized control trial. *J Natl Med Assoc*. Aug 2008;100(8):936-44. doi:10.1016/s0027-9684(15)31408-5

11.	Fakolade R, Adebayo SB, Anyanti J, Ankomah A. The impact of exposure to mass media campaigns and social support on levels and trends of HIV-related stigma and discrimination in Nigeria: tools for enhancing effective HIV prevention programmes. *J Biosoc Sci*. May 2010;42(3):395-407. doi:10.1017/S0021932009990538

12.	Ezedinachi EN, Ross MW, Meremiku M, et al. The impact of an intervention to change health workers' HIV/AIDS attitudes and knowledge in Nigeria: a controlled trial. *Public Health*. Mar 2002;116(2):106-12. doi:10.1038/sj.ph.1900834

13.	Nyblade L, Addo NA, Atuahene K, et al. Results from a difference-in-differences evaluation of health facility HIV and key population stigma-reduction interventions in Ghana. *J Int AIDS Soc*. Apr 2020;23(4):e25483. doi:10.1002/jia2.25483

14.	Pulerwitz J, Hui W, Arney J, Scott LM. Changing Gender Norms and Reducing HIV and Violence Risk Among Workers and Students in China. *J Health Commun*. Aug 2015;20(8):869-78. doi:10.1080/10810730.2015.1018573

15.	Uys L, Chirwa M, Kohi T, et al. Evaluation of a health setting-based stigma intervention in five African countries. *AIDS Patient Care STDS*. Dec 2009;23(12):1059-66. doi:10.1089/apc.2009.0085

16.	Arora S, Jyoti S, Chakravarty S. Effectiveness of an empowering programme on student nurses' understanding and beliefs about HIV/AIDS. *International Journal of Nursing Education*. 2014;6(1):88-92.

17.	Mak WW, Cheng SS, Law RW, Cheng WW, Chan F. Reducing HIV-related stigma among health-care professionals: a game-based experiential approach. *AIDS Care*. 2015;27(7):855-9. doi:10.1080/09540121.2015.1007113
